# Adjunctive Vasopressors in Patients with Septic Shock: Protocol for a Systematic Review and Meta-Analysis

**DOI:** 10.1101/2023.07.29.23293364

**Authors:** Seth R. Bauer, Patrick M. Wieruszewski, Brittany D. Bissell, Siddharth Dugar, Gretchen L. Sacha, Ryota Sato, Matthew T. Siuba, Mary Schleicher, Vidula Vachharajani, Yngve Falck-Ytter, Rebecca L. Morgan

**Author notes:** **Address correspondence to:** Seth R. Bauer, PharmD, FCCM, FCCP, Cleveland Clinic, Department of Pharmacy, Mail Code JJN1-284, 9500 Euclid Avenue, Cleveland, OH 44195. **Contributions:** Conceptualization: SRB, SD, VV. Methodology: SRB, PMW, BDB, SD, MS, YF-Y, RLM. Project administration: SRB, PMW, YF-Y, RLM. Supervision: VV, YF-Y, RLM. Writing – original draft: SRB. Writing – review and editing: SRB, PMW, BDB, SD, GLS, RS, MTS, MS, VV, YF-Y, RLM. SRB takes responsibility for the content of the manuscript. **Registration:** PROSPERO CRD42023427984.

## Abstract

**Background:** Over one-third of patients with septic shock have adjunctive vasopressors added to first-line vasopressors. However, no randomized trial has detected improved mortality with adjunctive vasopressors. Published systematic reviews and meta-analysis have sought to inform the use of adjunctive vasopressors, yet each published review has limitations that hinder its interpretation. This review aims to overcome the limitations of previous reviews by systematically synthesizing the direct evidence for adjunctive vasopressor therapy use in adult patients with septic shock.

**Methods:** We will conduct a systematic review and meta-analysis of randomized controlled trials evaluating adjunctive vasopressors (vasopressin analogues, angiotensin II, hydroxocobalamin, methylene blue, and catecholamine analogues) in adult patients with septic shock. Relevant studies will be identified through comprehensive searches of MEDLINE, Embase, CENTRAL, and reference lists of previous systematic reviews. Only randomized trials comparing adjunctive vasopressors (>75% of subjects on vasopressors at enrollment) to standard care vasopressors in adults with septic shock (>75% of subjects having septic shock) will be included. Titles and abstracts will be screened, full-text articles assessed for eligibility, and data extracted from included studies. Outcomes of interest include short-term mortality, intermediate-term mortality, kidney replacement therapy, digital/peripheral ischemia, and venous thromboembolism. Pairwise meta-analysis using a random-effects model will be utilized to estimate the risk ratio for the outcomes. Risk of bias will be adjudicated with the Cochrane Risk of Bias 2 tool, and GRADE will be used to rate the certainty of the body of evidence.

**Discussion:** Although adjunctive vasopressors are commonly used in patients with septic shock their effect on patient-important outcomes is unclear. This study is planned to use rigorous systematic review methodology, including strict adhere to established guidelines, in order to overcome limitations of previously-published reviews and inform clinical practice and treatment guidelines for the use of adjunctive vasopressors in adults with septic shock.

**Systematic review registration:** PROSPERO CRD4202327984

## Background

Worldwide, sepsis causes over 49 million yearly incident cases, leading to 11 million sepsis-related deaths (20% of all global deaths).^1^ Further, sepsis is the leading cause of death in United States hospitals.^2^ As such, interventions to improve sepsis mortality are urgently needed. Sepsis is defined as a dysregulated host response to an infection leading to life-threatening organ dysfunction.^3^ The most severe subset of sepsis is septic shock, characterized by cardiovascular failure due to vasodilation, resulting in hypotension that requires supportive care with vasopressors to maintain adequate blood pressure. Norepinephrine, a catecholamine, is established as the first-line vasopressor in patients with septic shock.^4^ However, adjunctive vasopressors, such as vasopressin or alternative catecholamines, are utilized in addition to norepinephrine in 33-42% of patients.^5,6^ Adjunctive vasopressors have rationale for their use due to different pharmacology than norepinephrine leading to alternative pathways for increasing blood pressure.

A number of large trials have evaluated adjunctive vasopressors in adults with septic shock.^7-10^ These trials most commonly compared vasopressin (and its analogues) or angiotensin II to either blinded norepinephrine or placebo, each with the continuation of open-label standard care vasopressors. Adjunctive vasopressors have been shown to increase blood pressure, reflected as either achievement of a mean arterial pressure (MAP) of ≥65 mm Hg or a decrease in open-label vasopressor dosage while maintaining MAP at goal. Yet, despite point estimates favoring adjunctive vasopressors, the treatment effect was small enough that no trial has detected improved mortality with an adjunctive vasopressor over standard care. Furthermore, only one trial has shown an improvement in a patient-important outcome with an adjunctive vasopressor.^8^ These findings have led to large practice variability in adjunctive vasopressor use both globally and within the United States.^5,11^

Previously published systematic reviews and meta-analyses have sought to inform the use of adjunctive vasopressors in patients with septic shock. These reviews are used broadly, including in treatment guidelines, to support the use of adjunctive vasopressors over norepinephrine monotherapy.^4,12-18^ But each of the published reviews has important limitations that hinder its interpretation. In particular, the review of vasopressin performed for the 2021 Surviving Sepsis Campaign International Guidelines is limited by misclassification of a before-after cohort study as a randomized controlled trial,^19^ inclusion of data from the same study twice (based on conference abstract and full text publications),^19,20^ inclusion of studies evaluating the vasopressor(s) of interest as first-line therapy instead of adjunctive therapy,^21,22^ and synthesis of indirect evidence from trials enrolling patients with the broader syndrome of vasodilatory/distributive shock (of which septic shock is one of several etiologies).^22^ Other published systematic reviews and meta-analyses have one or more of these limitations, which are important for three reasons. First, simultaneous synthesis of non-randomized studies evaluating asynchronous cohorts with randomized controlled trials in a pairwise meta-analysis can lead to confounding by selection bias, unexplained heterogeneity, and ambiguous results.^23,24^ Second, patients with septic shock have different hemodynamics and outcomes compared with patients with other vasodilatory/distributive shock etiologies (such as post-cardiovascular surgery).^25-27^ Lastly, and importantly, trials evaluating the vasopressor of interest as a first-line vasopressor are not directly applicable to the use of the vasopressor as an adjunctive agent. This systematic review and meta-analysis is designed to overcome these limitations, and inform bedside practice and treatment guidelines for the use of adjunctive vasopressors in adult patients with septic shock based on the direct evidence from randomized controlled trials. The review question is “In adult patients with septic shock, should we use an adjunctive vasopressor versus continuing standard care vasopressor(s)?”.

## Methods

The objectives of this study are, in adult patients with septic shock, to 1) estimate the overall effect of adjunctive vasopressors on patient-important outcomes, and 2) estimate the effect of unique adjunctive vasopressor drug class/mechanism of action, compared with standard care vasopressor(s), on patient-important outcomes. This protocol is registered with PROSPERO (CRD42023427984) and reported in alignment with the Preferred Reporting Items for Systematic Review and Meta-Analysis protocols (PRISMA-P) statement.^28^

### Eligibility criteria

This systematic review and meta-analysis will use the Population, Intervention, Comparator, Outcomes (PICO) framework to develop the clinical question.^29^ The population of interest in the studies for inclusion is adult patients with septic shock, as defined by the consensus definition in use at the time the study was conducted (whether the Sepsis-2 or Sepsis-3 definition).^3,30^ The intervention of interest is adjunctive vasopressor use, defined as a vasopressor drug added to (used as adjunctive therapy to) open-label standard care vasopressor(s). Adjunctive vasopressor drugs of interest are 1) vasopressin and its analogues pituitrin, selepressin, and terlipressin; 2) angiotensin II; 3) the nitric oxide pathway modulators methylene blue and hydroxocobalamin; and 4) the catecholamine (and derivatives thereof) vasopressors dopamine, epinephrine, and phenylephrine. The comparator in the studies for inclusion is standard care vasopressors, which may include a) placebo with continuation of open-label standard care vasopressors, or b) blinded comparator standard care vasopressor. Despite current recommendations for the use of norepinephrine as the first-line and standard care vasopressor for the treatment of adults with septic shock, previous iterations of the Surviving Sepsis Campaign International Guidelines have recommended either norepinephrine or dopamine as the standard care vasopressor.^4,31-34^ Further, the Surviving Sepsis Campaign International Guidelines suggestions or recommendations regarding adjunctive vasopressors have evolved over time. ^4,31-34^ Therefore, open-label or blinded infusion of norepinephrine or dopamine, with or without any adjunctive vasopressor infusion, will also be considered as standard care vasopressors for the purpose of this review. Outcomes of interest for this review are short-term mortality (≤30 days), intermediate-term mortality (60 days), kidney replacement therapy, digital/peripheral ischemia, and venous thromboembolism (including deep vein thrombosis and pulmonary embolism).

Studies will be included if they meet each of the following criteria: 1) randomized controlled trial; 2) evaluated hospitalized adult subjects (age 16 years or older); 3) at least 75% of study subjects had septic shock; 4) evaluated adjunctive vasopressor therapy, defined as at least 75% of study subjects receiving standard care vasopressor(s) at enrollment; and 5) utilized standard care vasopressor(s) as the comparator, inclusive of placebo with continuation of open-label standard care vasopressors. Studies published throughout the inclusive dates of the electronic searchers, whether reported in full manuscript or abstract form, will be considered for inclusion. Eligible studies will be included regardless of their intent with the intervention adjunctive vasopressor, whether to increase MAP to a goal level or to decrease open-label standard care vasopressor dose (while maintaining MAP at goal level). Studies will be excluded if they were published in a non-English language, had a cross-over design, or were missing all outcomes of interest.

### Information sources

We will search the electronic databases MEDLINE (via Ovid), Embase (via Ovid), and Cochrane Central Register of Controlled Trials (CENTRAL) from their inception to 7 June 2023. The search strategy for MEDLINE (via Ovid) is included in Table 1 and the search strategies for Embase (via Ovid) and CENTRAL are included in the Supplemental Material as eTable 1 and eTable 2, respectively. Searches will be restricted to studies published in the English language. We will also hand search the reference lists of included studies and relevant systematic reviews. Relevant systematic reviews will be identified by searching MEDLINE (via Ovid) with a similar search strategy as outlined in Table 1, with substitution of randomized controlled trial study design elements for systematic review study design elements. A preliminary search for relevant systematic reviews from 1946 to 6 February 2023 completed on 7 February 2023 identified 45 potential systematic reviews for evaluation. Furthermore, we will review regulatory agency documents for additional relevant study details, such as those provided on Drugs@FDA, and search for ongoing studies via querying ClinicalTrials.gov.

**Table 1.**
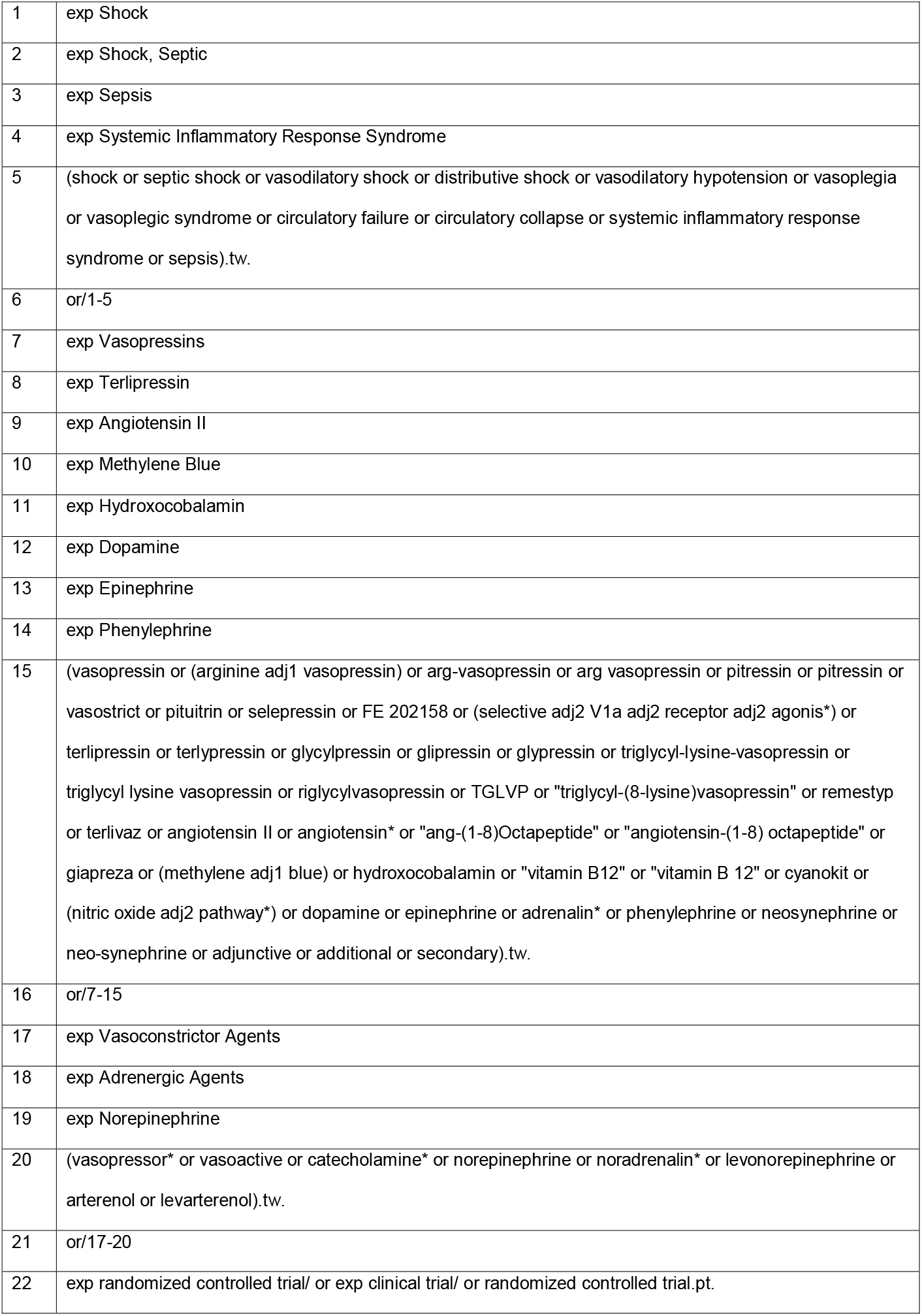
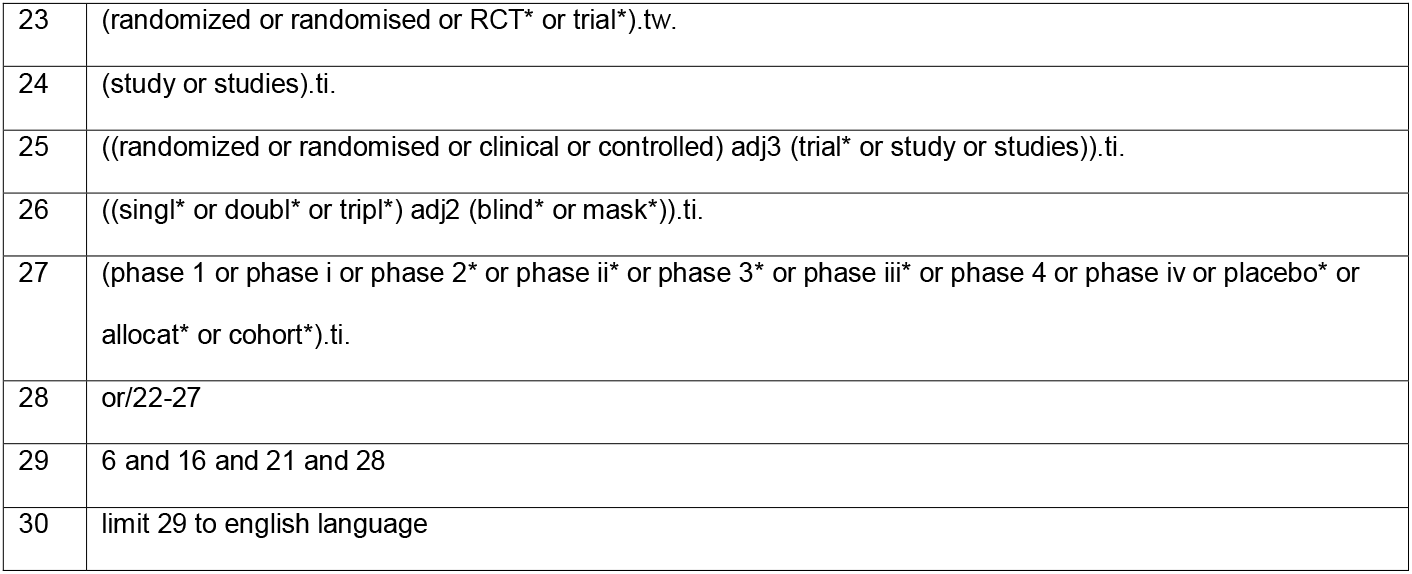
MEDLINE (via Ovid) Search Strategy.

**Table 2.**
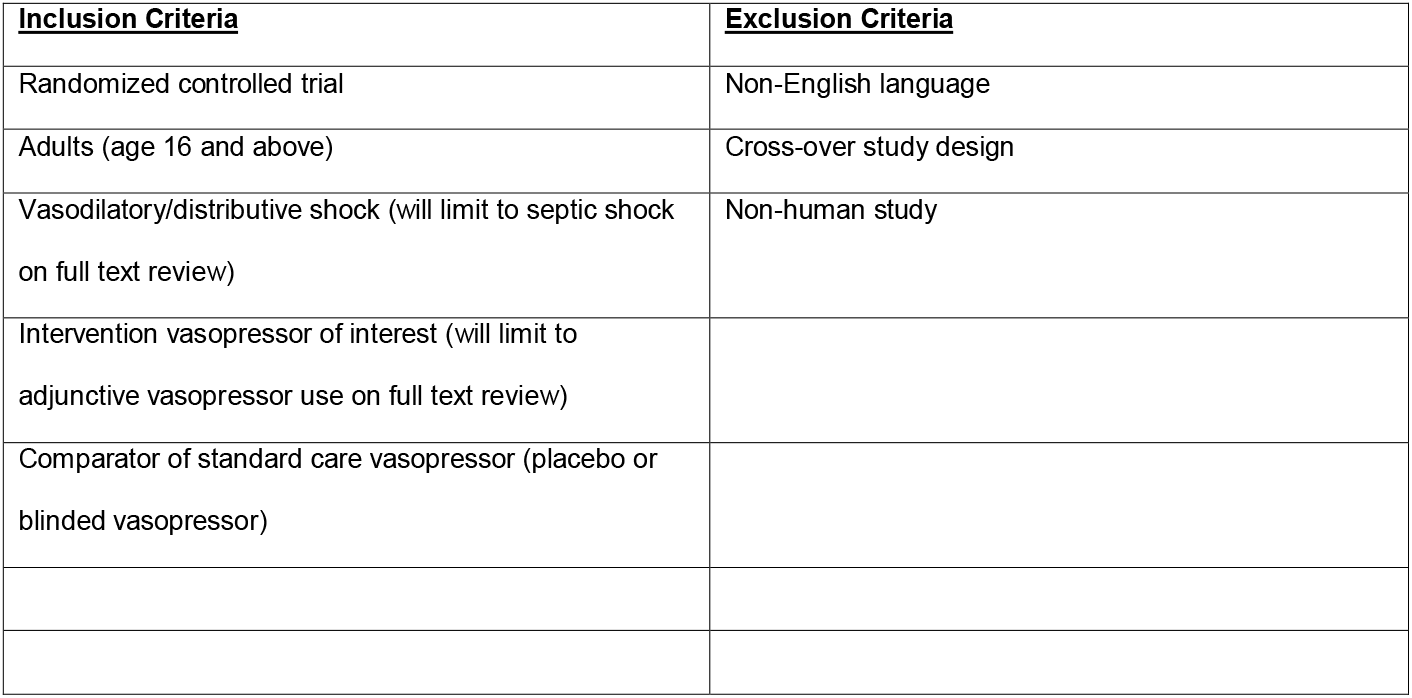
Title and abstract screening criteria.

### Study records and data items

The systematic review management software Covidence (Melbourne, Australia) will be utilized for study screening. The title and abstract of articles identified through electronic searchers will be imported into the software. Two review authors will independently assess titles and abstracts using standardized criteria. (Table 2) If the title and abstract do not provide sufficient information to determine potential for inclusion the full text will be retrieved. Two review authors will independently assess full text publications for inclusion using standardized criteria. (Table 3) Disagreements will be resolved by discussion and through consulting a third review author, if needed. The reasons for exclusion will be documented.

**Table 3.** Full text screening exclusion criteria.

| **<u>Exclusion Criteria</u>** |
| --- |
| Additional report of already addressed study |
| <75% of enrolled subjects with septic shock |
| <75% of enrolled subjects receiving vasopressors at enrollment |
| Included age $\leq 15$ years |
| Missing all outcomes of interest |
| Non-English language |
| Not randomized controlled trial |
| Ongoing study without results |
| Unable to retrieve full text |
| Wrong intervention |
| Wrong comparator |

A single review author will extract citation, population, intervention, comparator, and methodologic data from included studies using a piloted data extraction form. Multiple reports from the same study will be collated into one study record. We will extract citation information (authors, publication year, number of trial centers, country(ies), author-reported financial relationships and other potential conflicts of interest), and characteristics of the study population (number of included subjects, age, sex, percentage with White race, body weight, percentage with septic shock, percentage receiving baseline vasopressors, baseline standard care vasopressor [norepinephrine] dosage), intervention (drug, administration method, dosage, MAP goal), comparator (placebo, blinded standard care vasopressor administration method and dosage, open-label standard care vasopressor administration method and dosage), and outcomes (listed and defined below) using the electronic data capture tool REDCap (Vanderbilt University, Nashville, TN). Methodologic elements will also be extracted to enable adjudication for risk of bias. All outcome data will be extracted independently by two review authors, with discrepancies resolved by discussion. Any relevant retraction statements and errata for studies will be examined. The electronic data extraction instrument for included studies is in the Supplemental Material as eFigure 1. Extracted data will be compared to data from previously-published systematic reviews for consistencies. In the case of inconsistencies, study investigators will be contacted to confirm data. If investigators do not respond to the initial request they will be contacted once more. If no response is received after two attempts then the data extracted from the primary reference will be utilized.

### Outcomes

The main outcome for this study is short-term mortality, defined as all-cause mortality at ≤30 days or at intensive care unit discharge.^35^ Additional outcomes include a) intermediate-term mortality, defined as all-cause mortality at 60 days or at hospital discharge,^35^ b) kidney replacement therapy, defined as the need for continuous or intermittent kidney replacement therapy (“dialysis”) on one or more days, c) digital/peripheral ischemia, defined as evidence of excessive vasoconstriction leading to ischemia in the periphery or digits, and d) venous thromboembolism, defined as venous thrombotic and thromboembolic events. Short-term and intermediate-term mortality are considered patient-important outcomes in critically ill patients,^36^ and digital/peripheral ischemia and venous thromboembolism are recognized adverse effects of adjunctive vasopressors.^37^ Each of these outcomes of interest was deemed of “critical” importance by panel members for the 2021 Surviving Sepsis Campaign International Guidelines.^4^

### Risk of bias in individual studies

Two review authors will independently assess potential risk of bias for each outcome resulting from the trial design based on the Cochrane Risk of Bias 2 (RoB 2) tool for individually-randomized, parallel-group trials.^38^ In addition to published study manuscripts, trial protocols, registers, and regulatory documents will be utilized to assess potential risk of bias. The electronic data capture tool will use branching logic to ensure applicable signaling questions from RoB 2 are addressed based on responses to previous signaling questions. The data capture tool will also have embedded resources (such as those from riskofbias.info) to optimize accuracy and consistency of responses to signaling questions. Judgements of risk of bias will be justified in the data capture tool. The electronic risk of bias adjudication instrument for included studies is in the Supplemental Material as eFigure 2. Risk of bias adjudication discrepancies between review authors will be resolved through discussion. The risk of bias for each outcome of interest from each included trial will be summarized and displayed in figures for the outcome. These figures will be created with Review Manager 5.4 (Nordic Cochrane Centre, The Cochrane Collaboration).

### Data synthesis

Included studies determined to be sufficiently similar for grouping will be analyzed with pairwise meta-analysis. Treatment effect for each outcome (all dichotomous) will be assessed by calculating the risk ratio with 95% confidence interval between intervention (adjunctive vasopressor) and control (standard care vasopressors) groups. Meta-analysis will be conducted using a random-effects model with the Der Simonian and Laird inverse variance method to estimate the risk ratio with 95% confidence interval. If a single arm of a study has an observed zero count, continuity correction will be applied by adding 0.5 to the event count.^39^ If a study has zero counts observed in both study arms the study will be omitted from the meta-analysis because the risk ratio would be undefined. The results of the meta-analysis will be presented with a forest plot, including individual study intervention effect estimates (expressed as the risk ratio with 95% confidence interval) and study weight, the overall weighted average of the intervention effects estimated in the individual studies, and measures of heterogeneity. Heterogeneity among trials will be quantified with the I^2^ statistic, reflecting the percentage of total variability in effect estimates due to between-study heterogeneity rather than chance. A threshold for interpretation of the I^2^ statistic will not be utilized.^39^ Evidence for heterogeneity of the intervention effect beyond chance will be determined from the p-value of a chi-square test. A p-value threshold below 0.10 will be utilized to determine statistical significance.^39^ Heterogeneity will be further explored through subgroup and sensitivity analyses (described below). When possible, evidence for publication bias for each outcome will be assessed with funnel plots. Statistical analyses will be performed with RevMan 5.4 (Nordic Cochrane Centre, The Cochrane Collaboration).

### Subgroup and sensitivity analyses

We will carry out subgroup analyses to address the effect of 1) adjunctive vasopressor drug class/mechanism of action; 2) baseline shock severity, as assessed by norepinephrine-equivalent dosage above or below the overall median value from all studies; and 3) comparator standard care vasopressor (such as norepinephrine or dopamine). For the first subgroup analysis, adjunctive vasopressor drug class/mechanism of action will be grouped as a) vasopressin analogues, b) angiotensin II, c) methylene blue/hydroxocobalamin, or d) catecholamine analogues. For the second subgroup analysis, if norepinephrine-equivalent dosage (combining the dosage of all vasopressors together) is reported, this dosage will be primarily utilized. If norepinephrine-equivalent dosage is not reported then the norepinephrine dosage will be utilized, regardless of the reported presence or not of additional vasopressors. The norepinephrine-equivalent dosage in all included study participants will be primarily utilized; if this value is not reported then the norepinephrine dosage in the intervention arm will be utilized. In order to facilitate comparison between studies, norepinephrine-equivalent dosages expressed as mcg/min will be converted to mcg/kg/min by dividing by the reported mean (or median) body weight of patients in the study (or intervention arm, as appropriate). If body weight is not reported, norepinephrine-equivalent dosage in mcg/min will be divided by 75 kg to result in a mcg/kg/min dosage. If baseline norepinephrine-equivalent dosage or norepinephrine dosage is not reported, the dosage of the most-frequent baseline vasopressor will be converted to norepinephrine-equivalent dosage using a standardized formula.^40^ Norepinephrine dosages reported as median values (with range or interquartile range) in individual studies will have the mean value estimated prior to calculation of the overall median value for subgroup designation.^41^ We will also perform sensitivity analyses based on 1) risk of bias by comparing studies with an overall low risk of bias to those with some concerns/high risk of bias, and 2) inclusion of only trials evaluating non-catecholamine adjunctive vasopressors (because these agents increase blood pressure through a different mechanism than standard care catecholamine vasopressors, such as norepinephrine).

### Certainty in evidence

We will use the Grading of Recommendations Assessment, Development, and Evaluation (GRADE) system of rating the certainty (i.e., quality or confidence) of the body of evidence.^42^ The main findings of the review will be presented in a summary of findings table, which is a succinct, transparent, and informative summary of evidence in tabular format showing the quality of evidence and the extent of relative and absolute effects for each outcome. To ensure judgments are systematic and transparent, detailed quality assessments will be provided in evidence profiles with rationale for each factor that determines the quality of evidence for each outcome. GRADE approach will be utilized to make judgments about domains that decrease certainty in an estimate: risk of bias, inconsistency,^43^ indirectness,^44^ imprecision,^45^ and publication bias. The assessment of imprecision is based upon a threshold of a minimum clinically important difference, but a minimum clinically important difference for mortality in patients with septic shock, or critically ill patients in general, has not been established. Some have utilized a mortality absolute difference of 5% as the minimum clinically important difference based on review of the literature,^46^ while others have postulated a 1.5% minimum clinically important difference for calculation of statistical power in a trial.^47^ Due to this uncertainty we will consider both of these thresholds when adjudicating imprecision. We will use the GRADE approach to classify the certainty of evidence in one of four levels; high (we are very confident that the true effect lies close to that of the estimate of the effect), moderate (we are moderately confident in the estimate of effect; the true effect is likely to be close to the estimate of effect, but possibly substantially different), low (our confidence in the effect is limited: the true effect may be substantially different from the estimate of the effect), or very low (we have very little confidence in the effect estimate: the true effect is likely to be substantially different from the estimate of effect). We will use GRADEpro GDT software to facilitate certainty of evidence assessment.^48^

## Discussion

A number of systematic reviews and meta-analyses have been published evaluating adjunctive vasopressors in patients with septic shock. However, each published review has important limitations that hinders its interpretation. This study is planned to use rigorous systematic review methodology, including strict adhere to established guidelines, in order to overcome limitations of previously-published analyses. The study report will adhere to the PRISMA 2020 statement,^49^ and will be submitted for publication in a peer-reviewed journal with consideration of open access publication in order to increase dissemination. The report will inform bedside practice and treatment guidelines for the use of adjunctive vasopressors in adult patients with septic shock.

## Data Availability

All data produced in the present work are contained in the manuscript

## References

1. Rudd KE, Johnson SC, Agesa KM, et al. Global, regional, and national sepsis incidence and mortality, 1990–2017: analysis for the Global Burden of Disease Study. Lancet. 2020;395(10219):200–200.

2. Liu V, Escobar GJ, Greene JD, et al. Hospital Deaths in Patients With Sepsis From 2 Independent Cohorts. JAMA. 2014;312(1):90–90.

3. Singer M, Deutschman CS, Seymour CW, et al. The Third International Consensus Definitions for Sepsis and Septic Shock (Sepsis-3). JAMA. 2016;315(8):801–801.

4. Evans L, Rhodes A, Alhazzani W, et al. Surviving Sepsis Campaign: International Guidelines for Management of Sepsis and Septic Shock 2021. Crit Care Med. 2021;49(11):e1063–e1143.

5. Vail EA, Gershengorn HB, Hua M, Walkey AJ, Wunsch H. Epidemiology of Vasopressin Use for Adults with Septic Shock. Ann Am Thorac Soc. 2016;13(10):1760–1760.

6. Bosch NA, Teja B, Wunsch H, Walkey AJ. Practice Patterns in the Initiation of Secondary Vasopressors and Adjunctive Corticosteroids during Septic Shock in the United States. Ann Am Thorac Soc. 2021;18(12):2049–2049.

7. Russell JA, Walley KR, Singer J, et al. Vasopressin versus norepinephrine infusion in patients with septic shock. N Engl J Med. 2008;358(9):877–877.

8. Gordon AC, Mason AJ, Thirunavukkarasu N, et al. Effect of Early Vasopressin vs Norepinephrine on Kidney Failure in Patients With Septic Shock: The VANISH Randomized Clinical Trial. JAMA. 2016;316(5):509–509.

9. Laterre PF, Berry SM, Blemings A, et al. Effect of Selepressin vs Placebo on Ventilator- and Vasopressor-Free Days in Patients With Septic Shock: The SEPSIS-ACT Randomized Clinical Trial. JAMA. 2019;322(15):1476–1476.

10. Khanna A, English SW, Wang XS, et al. Angiotensin II for the Treatment of Vasodilatory Shock. N Engl J Med. 2017;377(5):419–419.

11. Abril MK, Khanna AK, Kroll S, McNamara C, Handisides D, Busse LW. Regional differences in the treatment of refractory vasodilatory shock using Angiotensin II in High Output Shock (ATHOS-3) data. J Crit Care. 2019;50:188–194.

12. Honarmand K, Um KJ, Belley-Cote EP, et al. Canadian Critical Care Society clinical practice guideline: The use of vasopressin and vasopressin analogues in critically ill adults with distributive shock. Can J Anaesth. 2020;67(3):369–369.

13. Avni T, Lador A, Lev S, Leibovici L, Paul M, Grossman A. Vasopressors for the Treatment of Septic Shock: Systematic Review and Meta-Analysis. PLoS One. 2015;10(8):e0129305.

14. Belletti A, Benedetto U, Biondi-Zoccai G, et al. The effect of vasoactive drugs on mortality in patients with severe sepsis and septic shock. A network meta-analysis of randomized trials. J Crit Care. 2017;37:91–98.

15. Zhong L, Ji X-W, Wang H-L, Zhao G-M, Zhou Q, Xie B. Non-catecholamine vasopressors in the treatment of adult patients with septic shock—evidence from meta-analysis and trial sequential analysis of randomized clinical trials. J Intensive Care. 2020;8(1):83.

16. Nagendran M, Russell JA, Walley KR, et al. Vasopressin in septic shock: an individual patient data meta-analysis of randomised controlled trials. Intensive Care Med. 2019;45(6):844–844.

17. Pasin L, Umbrello M, Greco T, et al. Methylene blue as a vasopressor: a meta-analysis of randomised trials. Crit Care Resusc. 2013;15(1):42–42.

18. McIntyre WF, Um KJ, Alhazzani W, et al. Association of Vasopressin Plus Catecholamine Vasopressors vs Catecholamines Alone With Atrial Fibrillation in Patients With Distributive Shock: A Systematic Review and Meta-analysis. JAMA. 2018;319(18):1889–1889.

19. Hammond DA, Ficek OA, Painter JT, et al. Prospective Open-label Trial of Early Concomitant Vasopressin and Norepinephrine Therapy versus Initial Norepinephrine Monotherapy in Septic Shock. Pharmacotherapy. 2018;38(5):531–531.

20. Clem O, Painter J, Cullen J, et al. Norepinephrine and Vasopressin vs Norepinephrine Alone for Septic Shock: Randomized Controlled Trial. Crit Care Med. 2016;44(12):413.

21. Morelli A, Ertmer C, Rehberg S, et al. Continuous terlipressin versus vasopressin infusion in septic shock (TERLIVAP): a randomized, controlled pilot study. Crit Care. 2009;13(4):R130.

22. Hajjar LA, Vincent JL, Barbosa Gomes Galas FR, et al. Vasopressin versus Norepinephrine in Patients with Vasoplegic Shock after Cardiac Surgery: The VANCS Randomized Controlled Trial. Anesthesiology. 2017;126(1):85–85.

23. Valentine JC, Thompson SG. Issues relating to confounding and meta-analysis when including non-randomized studies in systematic reviews on the effects of interventions. Res Synth Methods. 2013;4(1):26–26.

24. Sarri G, Patorno E, Yuan H, et al. Framework for the synthesis of non-randomised studies and randomised controlled trials: a guidance on conducting a systematic review and meta-analysis for healthcare decision making. BMJ Evid Based Med. 2022;27(2):109–109.

25. Geri G, Vignon P, Aubry A, et al. Cardiovascular clusters in septic shock combining clinical and echocardiographic parameters: a post hoc analysis. Intensive Care Med. 2019;45(5):657–657.

26. Lambden S, Creagh-Brown BC, Hunt J, Summers C, Forni LG. Definitions and pathophysiology of vasoplegic shock. Crit Care. 2018;22(1):174.

27. Levy B, Fritz C, Tahon E, Jacquot A, Auchet T, Kimmoun A. Vasoplegia treatments: the past, the present, and the future. Crit Care. 2018;22(1):52.

28. Shamseer L, Moher D, Clarke M, et al. Preferred reporting items for systematic review and meta-analysis protocols (PRISMA-P) 2015: elaboration and explanation. BMJ. 2015;350:g7647.

29. Thomas J, Kneale D, McKenzie JE, Brennan SE, Bhaumik S. Chapter 2: Determining the scope of the review and the questions it will address. In: Higgins JPT, Thomas J, Chandler J, et al., eds. Cochrane Handbook for Systematic Reviews of Interventions. 6.3 (updated February 2022) ed: Cochrane; 2022.

30. Levy MM, Fink MP, Marshall JC, et al. 2001 SCCM/ESICM/ACCP/ATS/SIS International Sepsis Definitions Conference. Crit Care Med. 2003;31(4):1250–1250.

31. Dellinger RP, Carlet JM, Masur H, et al. Surviving Sepsis Campaign guidelines for management of severe sepsis and septic shock. Crit Care Med. 2004;32(3):858–858.

32. Dellinger RP, Levy MM, Carlet JM, et al. Surviving Sepsis Campaign: international guidelines for management of severe sepsis and septic shock: 2008. Crit Care Med. 2008;36(1):296–296.

33. Dellinger RP, Levy MM, Rhodes A, et al. Surviving sepsis campaign: international guidelines for management of severe sepsis and septic shock: 2012. Crit Care Med. 2013;41(2):580–580.

34. Rhodes A, Evans LE, Alhazzani W, et al. Surviving Sepsis Campaign: International Guidelines for Management of Sepsis and Septic Shock: 2016. Crit Care Med. 2017;45(3):486–486.

35. Friedrich JO, Harhay MO, Angus DC, et al. Mortality As a Measure of Treatment Effect in Clinical Trials Recruiting Critically Ill Patients. Crit Care Med. 2023;51(2):222–222.

36. Gaudry S, Messika J, Ricard J-D, et al. Patient-important outcomes in randomized controlled trials in critically ill patients: a systematic review. Ann Intensive Care. 2017;7(1):28.

37. Bauer SR, Sacha GL, Lam SW. Safe Use of Vasopressin and Angiotensin II for Patients with Circulatory Shock. Pharmacotherapy. 2018;38(8):851–851.

38. Sterne JAC, Savoviþ J, Page MJ, et al. RoB 2: a revised tool for assessing risk of bias in randomised trials. BMJ. 2019;366:l4898.

39. Deeks JJ, Higgins JPT, Altman DJ. Chapter 10: Analysing data and undertaking meta-analyses. In: Higgins JPT, Thomas J, Chandler J, et al., eds. Cochrane Handbook for Systematic Reviews of Interventions. Vol 6.3 (updated February 2022): Cochrane; 2022.

40. Goradia S, Sardaneh AA, Narayan SW, Penm J, Patanwala AE. Vasopressor dose equivalence: A scoping review and suggested formula. J Crit Care. 2021;61:233–240.

41. Wan X, Wang W, Liu J, Tong T. Estimating the sample mean and standard deviation from the sample size, median, range and/or interquartile range. BMC Med Res Methodol. 2014;14:135.

42. Guyatt G, Oxman AD, Akl EA, et al. GRADE guidelines: 1. Introduction-GRADE evidence profiles and summary of findings tables. J Clin Epidemiol. 2011;64(4):383–383.

43. Guyatt GH, Oxman AD, Kunz R, et al. GRADE guidelines: 7. Rating the quality of evidence - inconsistency. J Clin Epidemiol. 2011;64(12):1294–1294.

44. Guyatt GH, Oxman AD, Kunz R, et al. GRADE guidelines: 8. Rating the quality of evidence - indirectness. J Clin Epidemiol. 2011;64(12):1303–1303.

45. Guyatt GH, Oxman AD, Kunz R, et al. GRADE guidelines 6. Rating the quality of evidence - imprecision. J Clin Epidemiol. 2011;64(12):1283–1283.

46. Abrams D, Montesi SB, Moore SKL, et al. Powering Bias and Clinically Important Treatment Effects in Randomized Trials of Critical Illness. Crit Care Med. 2020;48(12):1710–1710.

47. Young P, Arabi Y, Bagshaw S, et al. Protocol and statistical analysis plan for the mega randomised registry trial research program comparing conservative versus liberal oxygenation targets in adults receiving unplanned invasive mechanical ventilation in the ICU (Mega-ROX). Crit Care Resusc. 2022;24(2):137–137.

48. GRADEpro GDT: GRADEpro Guideline Development Tool [Software]. McMaster University and Evidence Prime, 2022. Available from gradepro.org.

49. Page MJ, McKenzie JE, Bossuyt PM, et al. The PRISMA 2020 statement: an updated guideline for reporting systematic reviews. BMJ. 2021;372:n71.

