## Supplemental Material for "Adjunctive Vasopressors in Patients with Septic Shock: Protocol for a Systematic Review and Meta-Analysis"

**Table of Contents**

eTable 1. Embase (via Ovid) Search Strategy

eTable 2. Cochrane Central Register of Controlled Trials (CENTRAL) Search Strategy

eFigure 1. Electronic data extraction instrument for included studies

eFigure 2. Electronic risk of bias adjudication instrument for included studies

**eTable 1**. Embase (via Ovid) Search Strategy

| | |
|---|---|
| 1 | exp shock/ |
| 2 | exp septic shock/ |
| 3 | exp sepsis/ |
| 4 | exp systemic inflammatory response syndrome/ |
| 5 | (shock or septic shock or vasodilatory shock or distributive shock or vasodilatory hypotension or vasoplegia or vasoplegic syndrome or circulatory failure or circulatory collapse or systemic inflammatory response syndrome or sepsis).tw. |
| 6 | or/1-5 |
| 7 | exp vasopressin/ |
| 8 | exp terlipressin/ |
| 9 | exp angiotensin II/ |
| 10 | exp methylene blue/ |
| 11 | exp hydroxocobalamin/ |
| 12 | exp dopamine/ |
| 13 | exp epinephrine/ |
| 14 | exp phenylephrine/ |
| 15 | (vasopressin or (arginine adj1 vasopressin) or arg-vasopressin or arg vasopressin or pitressin or vasostrict or pituitrin or selepressin or FE 202158 or (selective adj2 V1a adj2 receptor adj2 agonis*) or terlipressin or terlypressin or glycylpressin or glipressin or glypressin or triglycyl-lysine-vasopressin or triglycyl lysine vasopressin or riglycylvasopressin or TGLVP or "triglycyl-(8-lysine)vasopressin" or remestyp or terlivaz or angiotensin II or angiotensin* or "ang-(1-8)Octapeptide" or "angiotensin-(1-8) octapeptide" or giapreza or (methylene adj1 blue) or hydroxocobalamin or "vitamin B12" or "vitamin B 12" or cyanokit or (nitric oxide adj2 pathway*) or dopamine or epinephrine or adrenalin* or phenylephrine or neosynephrine or neo-synephrine or adjunctive or additional or secondary).tw. |
| 16 | or/7-15 |
| 17 | exp vasoconstrictor agent/ |
| 18 | exp Adrenergic Agents/ |
| 19 | exp norepinephrine/ |
| 20 | (vasopressor* or vasoactive or catecholamine* or norepinephrine or noradrenalin* or levonorepinephrine or arterenol or levarterenol).tw. |
| 21 | or/17-20 |

| | |
|---|---|
| 22 | exp randomized controlled trial/ or exp clinical trial/ |
| 23 | (randomized or randomised or RCT* or trial*).tw. |
| 24 | (study or studies).ti. |
| 25 | ((randomized or randomised or clinical or controlled) adj3 (trial* or study or studies)).ti. |
| 26 | ((singl* or doubl* or tripl*) adj2 (blind* or mask*)).ti. |
| 27 | (phase 1 or phase i or phase 2* or phase ii* or phase 3* or phase iii* or phase 4 or phase iv or placebo* or allocat* or cohort*).ti. |
| 28 | or/22-27 |
| 29 | 6 and 16 and 21 and 28 |
| 30 | limit 29 to english language |

**eTable 2**. Cochrane Central Register of Controlled Trials (CENTRAL) Search Strategy

| | |
|---|---|
| 1 | [mh Shock] |
| 2 | [mh "Shock, Septic"] |
| 3 | [mh Sepsis] |
| 4 | [mh "Systemic Inflammatory Response Syndrome"] |
| 5 | (shock or septic shock or vasodilatory shock or distributive shock or vasodilatory hypotension or vasoplegia or vasoplegic syndrome or circulatory failure or circulatory collapse or systemic inflammatory response syndrome or sepsis):ti,ab |
| 6 | [OR #1-#5] |
| 7 | [mh Vasopressins] |
| 8 | [mh Terlipressin] |
| 9 | [mh "Angiotensin II"] |
| 10 | [mh "Methylene Blue"] |
| 11 | [mh Hydroxocobalamin] |
| 12 | [mh Dopamine] |
| 13 | [mh Epinephrine] |
| 14 | [mh Phenylephrine] |
| 15 | (vasopressin or (arginine NEAR/1 vasopressin) or arg-vasopressin or arg vasopressin or pitressin or vasostrict or pituitrin or selepressin or FE 202158 or (selective NEAR/2 V1a NEAR/2 receptor NEAR/2 agonis*) or terlipressin or terlypressin or glycylpressin or glipressin or glypressin or triglycyl-lysine-vasopressin or triglycyl lysine vasopressin or riglycylvasopressin or TGLVP or "triglycyl-(8-lysine)vasopressin" or remestyp or terlivaz or angiotensin II or angiotensin* or "ang-(1-8)Octapeptide" or "angiotensin-(1-8) octapeptide" or giapreza or (methylene NEAR/1 blue) or hydroxocobalamin or "vitamin B12" or "vitamin B 12" or cyanokit or (nitric oxide NEAR/2 pathway*) or dopamine or epinephrine or adrenalin* or phenylephrine or neosynephrine or neo-synephrine or adjunctive or additional or secondary):ti,ab |
| 16 | [OR #7-#15] |
| 17 | [mh "Vasoconstrictor Agents"] |
| 18 | [mh "Adrenergic Agents"] |
| 19 | [mh Norepinephrine] |
| 20 | (vasopressor* or vasoactive or catecholamine* or norepinephrine or noradrenalin* or levonorepinephrine or arterenol or levarterenol):ti,ab |

| | |
|---|---|
| 21 | [OR #17-#20] |
| 22 | [mh "randomized controlled trial"] or [mh "clinical trial"] or randomized controlled trial:pt |
| 23 | (randomized or randomised or RCT* or trial*):ti,ab |
| 24 | (study or studies):ti |
| 25 | ((randomized or randomised or clinical or controlled) NEAR/3 (trial* or study or studies)):ti |
| 26 | ((singl* or doubl* or tripl*) NEAR/2 (blind* or mask*)):ti |
| 27 | (phase 1 or phase i or phase 2* or phase ii* or phase 3* or phase iii* or phase 4 or phase iv or placebo* or allocat* or cohort*):ti |
| 28 | [OR #22-#27] |
| 29 | #6 AND #16 AND #21 AND #28 in Trials |

**eFigure 1**. Electronic data extraction instrument for included studies

### Citation, population, intervention, comparator, and methodologic data

| | |
|---|---|
| First Author and Year | __________________ |
| Upload primary study report utilized for this data extraction | |
| Any additional study reports to upload? | ◯ Yes<br>◯ No |
| Number of additional reports | ◯ 1<br>◯ 2<br>◯ 3 |
| Upload additional study report utilized for this data extraction | |
| Upload second additional study report utilized for this data extraction | |
| Upload third additional study report utilized for this data extraction | |

| **Study Characteristics** | |
|---|---|
| Study Report Type | ☐ Manuscript<br>☐ Abstract<br>☐ Regulatory documents<br>☐ Trial register<br>(Check all that apply) |
| Study Confirmed as Meeting Inclusion Criteria? | ◯ Yes<br>◯ No |
| Trial Funding Mechanism | ◯ Government<br>◯ Industry<br>◯ Foundation<br>◯ Institutional<br>◯ Multiple |
| Multiple Funding Mechanisms | __________________<br>(List types (government, industry, foundation, institutional)) |
| Single or Multi-Center? | ◯ Multiple<br>◯ Single |
| Number of Study Centers | __________________ |

| | |
|---|---|
| Country | ◯ Multiple<br>◯ USA<br>◯ Austria<br>◯ Brazil<br>◯ China<br>◯ Czech Republic<br>◯ Egypt<br>◯ England<br>◯ India<br>◯ Iran<br>◯ Italy<br>◯ Russia<br>◯ Other |
| Other Country | ________________ |
| Setting | ◯ ICU<br>◯ Other |
| Other Setting | ________________ |
| Study Enrollment Start | ________________<br>(Year only) |
| Study Enrollment End | ________________<br>(Year only) |
| Outcomes Assessed | ☐ Short-term mortality (28-30d, ICU)<br>☐ Intermediate-term mortality (60d, Hospital)<br>☐ Kidney replacement therapy<br>☐ Digital/peripheral ischemia<br>☐ Venous thromboembolism<br>(Check all that apply) |
| Reminder: Complete risk of bias form for each outcome assessed | |
| Authors' financial relationship and other potential conflicts of interest | ________________<br>(Copy/Paste from report) |

**Patient Characteristics**

| | |
|---|---|
| Subjects Randomized in INTERVENTION Arm | ________________ |
| Subjects Randomized in COMPARATOR Arm | ________________ |
| Age central tendency and variance type | ◯ mean (SD)<br>◯ median (IQR) |

| | |
|---|---|
| Age, mean | ______________________ (Years. Prefer value in all included patients, if not available, collect value from intervention group. ) |
| Age, standard deviation | ______________________ (Years. Prefer value in all included patients, if not available, collect value from intervention group. ) |
| Age, median | ______________________ (Years. Prefer value in all included patients, if not available, collect value from intervention group. ) |
| Age, interquartile range | ______________________ (Years. Prefer value in all included patients, if not available, collect value from intervention group. ) |
| Female, % | ______________________ (Percentage from 0 to 100. Prefer value in all included patients, if not available, collect value from intervention group. ) |
| White race, % | ______________________ (Percentage from 0 to 100. Prefer value in all included patients, if not available, collect value from intervention group. ) |
| Body weight reported? | ◯ Yes<br>◯ No |
| Weight, kg | ______________________ (Either mean or median overall sample (or intervention) body weight.) |
| Septic shock, % | ______________________ (Overall study cohort; Percentage from 0 to 100) |
| Baseline Vasopressors at Enrollment, % | ______________________ (Overall study cohort; Percentage from 0 to 100) |
| Baseline Vasopressors at Enrollment | ☐ Norepinephrine<br>☐ Dopamine<br>☐ Epinephrine<br>☐ Phenylephrine<br>☐ Vasopressin<br>(Check all that apply) |

| | |
|---|---|
| Baseline NE dose type | ◯ Norepinephrine-equivalent dose (NEQ)<br>◯ Norepinephrine dose<br>◯ Not applicable<br>(NEQ preferred, if available) |
| Baseline NE dose units | ◯ mcg/min<br>◯ mcg/kg/min |
| Baseline NE dose central tendency and variance type | ◯ mean (SD)<br>◯ median (IQR) |
| Baseline NE dose, mean | ____________________<br>(Prefer value in all included patients, if not available, collect value from intervention group. If only receiving dopamine denote in field) |
| Baseline NE dose, standard deviation | ____________________ |
| Baseline NE dose, median | ____________________<br>(Prefer value in all included patients, if not available, collect value from intervention group. If only receiving dopamine denote in field) |
| Baseline NE dose, interquartile range | ____________________ |
| Baseline APACHE II score central tendency and variance type | ◯ mean (SD)<br>◯ median (IQR) |
| Baseline APACHE II, mean | ____________________<br>(Prefer value in all included patients, if not available, collect value from intervention group) |
| Baseline APACHE II, standard deviation | ____________________<br>(Prefer value in all included patients, if not available, collect value from intervention group) |
| Baseline APACHE II, median | ____________________ |
| Baseline APACHE II, IQR | ____________________ |

#### Intervention Characteristics

| | |
|---|---|
| MAP target | ◯ >=65<br>◯ >=70<br>◯ 65-75<br>◯ Other<br>(mm Hg) |
| Other MAP goal (mm Hg) | ________________________ |
| Adjunctive vasopressor group | ◯ Vasopressin analogue (vasopressin, terlipressin, selepressin, pituitrin)<br>◯ Angiotensin II<br>◯ Methylene blue and hydroxocobalamin<br>◯ Catecholamine vasopressor (dopamine, epinephrine, phenylephrine) |
| Specific Adjunctive Vasopressor | ◯ Vasopressin<br>◯ Terlipressin<br>◯ Selepressin<br>◯ Pituitrin<br>◯ Angiotensin II<br>◯ Methylene Blue<br>◯ Hydroxocobalamin<br>◯ Epinephrine<br>◯ Phenylephrine<br>◯ Dopamine |
| Relevant Study Co-Interventions? | ◯ Yes<br>◯ No |
| Describe co-interventions in detail (name, dose, duration) | ________________________<br>(Copy/Paste from report) |
| Administration type | ◯ IV infusion<br>◯ IV bolus<br>◯ IV bolus + infusion |
| Infusion starting dose | ________________________<br>(Include units) |
| Infusion minimum dose | ________________________<br>(Include units) |
| Infusion maximum dose | ________________________<br>(Include units) |
| IV bolus dose | ________________________<br>(Include units) |
| IV bolus frequency | ◯ Once<br>◯ Scheduled frequency |

| | |
|---|---|
| Scheduled frequency | ____________________ (Eg: q4h, q6h, etc.) |
| Hours from shock onset to enrollment | ____________________ (Mean (SD); if median (IQR or range) reported denote in field) |

#### Comparator Characteristics

| | |
|---|---|
| Comparator type | ◯ Placebo with open-label standard care vasopressor<br>◯ Blinded standard care vasopressor<br>◯ Only open-label standard care vasopressor |
| Blinded standard care vasopressor starting dose | ____________________ (Include units) |
| Blinded standard care vasopressor minimum dose | ____________________ (Include units) |
| Blinded standard care vasopressor maximum dose | ____________________ (Include units) |

#### Outcomes

| | |
|---|---|
| Short-term mortality timepoint | ◯ 28-day<br>◯ 30-day<br>◯ ICU |
| Short-term mortality in INTERVENTION group, numerator | ____________________ (Numerator) |
| Short-term mortality in INTERVENTION group, denominator | ____________________ (Denominator) |
| Short-term mortality in COMPARATOR group, numerator | ____________________ (Numerator) |
| Short-term mortality in COMPARATOR group, denominator | ____________________ (Denominator) |
| Intermediate-term mortality timepoint | ◯ 60-day<br>◯ Hospital |
| Intermediate-term mortality in INTERVENTION group, numerator | ____________________ (Numerator) |

| | |
|---|---|
| Intermediate-term mortality in INTERVENTION group, denominator | ______________________<br>(Denominator) |
| Intermediate-term mortality in COMPARATOR group, numerator | ______________________<br>(Numerator) |
| Intermediate-term mortality in COMPARATOR group, denominator | ______________________<br>(Denominator) |
| Kidney-replacement therapy in INTERVENTION group, numerator | ______________________<br>(Numerator) |
| Kidney-replacement therapy in INTERVENTION group, denominator | ______________________<br>(Denominator) |
| Kidney-replacement therapy in COMPARATOR group, numerator | ______________________<br>(Numerator) |
| Kidney-replacement therapy in COMPARATOR group, denominator | ______________________<br>(Denominator) |
| Digital/peripheral ischemia in INTERVENTION group, numerator | ______________________<br>(Numerator) |
| Digital/peripheral ischemia in INTERVENTION group, denominator | ______________________<br>(Denominator) |
| Digital/peripheral ischemia in COMPARATOR group, numerator | ______________________<br>(Numerator) |
| Digital/peripheral ischemia in COMPARATOR group, denominator | ______________________<br>(Denominator) |
| Venous thromboembolism in INTERVENTION group, numerator | ______________________<br>(Numerator. Includes both DVT and PE if reported separately) |
| Venous thromboembolism in INTERVENTION group, denominator | ______________________<br>(Denominator. Includes both DVT and PE if reported separately) |

| | |
|---|---|
| Venous thromboembolism in COMPARATOR group, numerator | ______________________________ (Numerator. Includes both DVT and PE if reported separately) |
| Venous thromboembolism in COMPARATOR group, denominator | ______________________________ (Denominator. Includes both DVT and PE if reported separately) |
| Include additional study notes or comments here | ______________________________ |

**eFigure 2.** Electronic risk of bias adjudication instrument for included studies

### Risk of bias assessment

First Author and Year ______________________________

See attached "Cribsheet" for brief explanation of each signaling question in RoB 2

[Attachment: "20190814_RoB_2.0_cribsheet_parallel_trial .pdf"]

#### 1. Bias arising from the randomization process

| | Yes (Y) | Probably Yes (PY) | No (N) | Probably No (PN) | No Information (NI) |
|---|---|---|---|---|---|
| 1.1 Was the allocation sequence random? | ○ | ○ | ○ | ○ | ○ |
| 1.2 Was the allocation sequence concealed until participants were enrolled and assigned to interventions? | ○ | ○ | ○ | ○ | ○ |
| 1.3 Did baseline differences between intervention groups suggest a problem with the randomization process? | ○ | ○ | ○ | ○ | ○ |

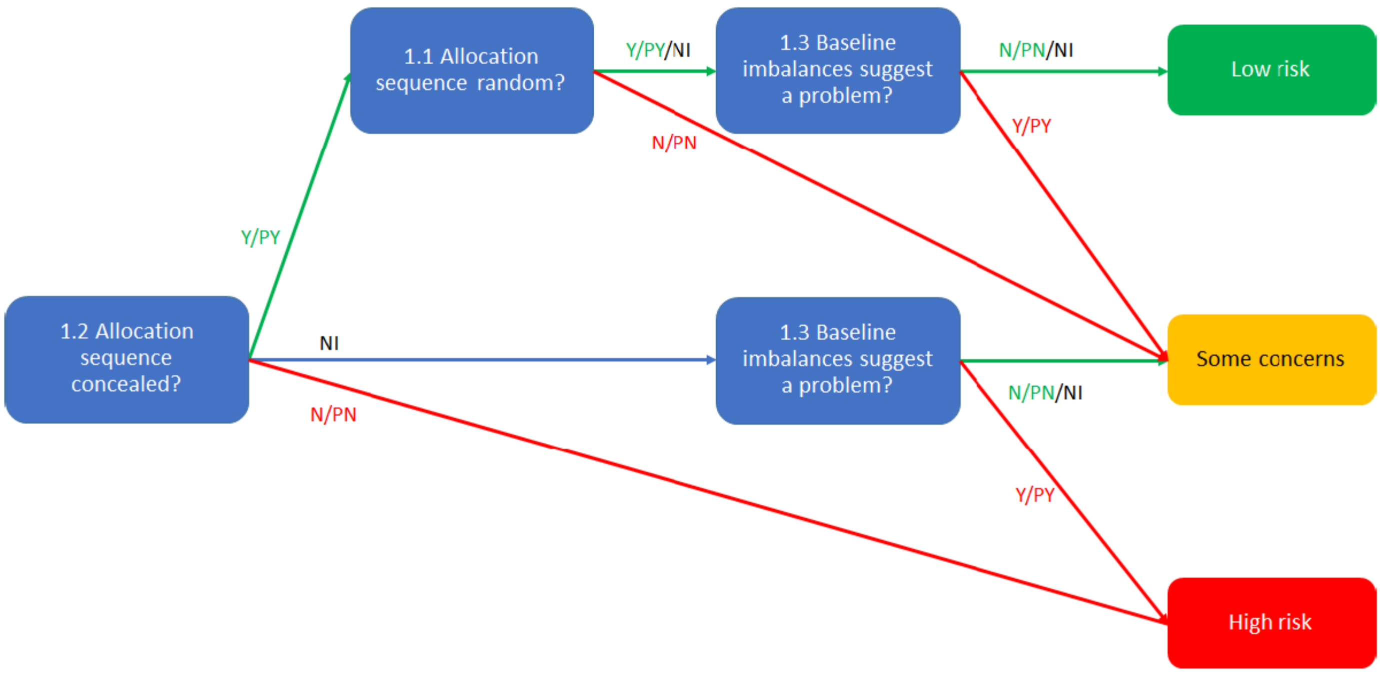


1. Risk of bias judgment: Randomization

○ Low
○ High
○ Some concerns
(Use above image, if needed, to assist with adjudication based on signaling question response and corresponding bias risk)

Include justification for judgment related to randomization process here

(Includes direct quotations from study report text, summary of information from a trial report, etc.)

#### 2. Bias due to deviations from intended interventions

| | Yes (Y) | Probably Yes (PY) | No (N) | Probably No (PN) | No Information (NI) |
|---|---|---|---|---|---|
| 2.1 Were participants aware of their assigned intervention during the trial? | ○ | ○ | ○ | ○ | ○ |
| 2.2 Were carers and people delivering the interventions aware of participants' assigned intervention during the trial? | ○ | ○ | ○ | ○ | ○ |
| 2.3 Were there deviations from the intended intervention that arose because of the trial context? | ○ | ○ | ○ | ○ | ○ |
| 2.4 Were these deviations likely to have affected the outcome? | ○ | ○ | ○ | ○ | ○ |
| 2.5 Were these deviations from intended intervention balanced between groups? | ○ | ○ | ○ | ○ | ○ |
| 2.6 Was an appropriate analysis used to estimate the effect of assignment to intervention? | ○ | ○ | ○ | ○ | ○ |
| 2.7 Was there potential for a substantial impact (on the result) of the failure to analyse participants in the group to which they were randomized? | ○ | ○ | ○ | ○ | ○ |

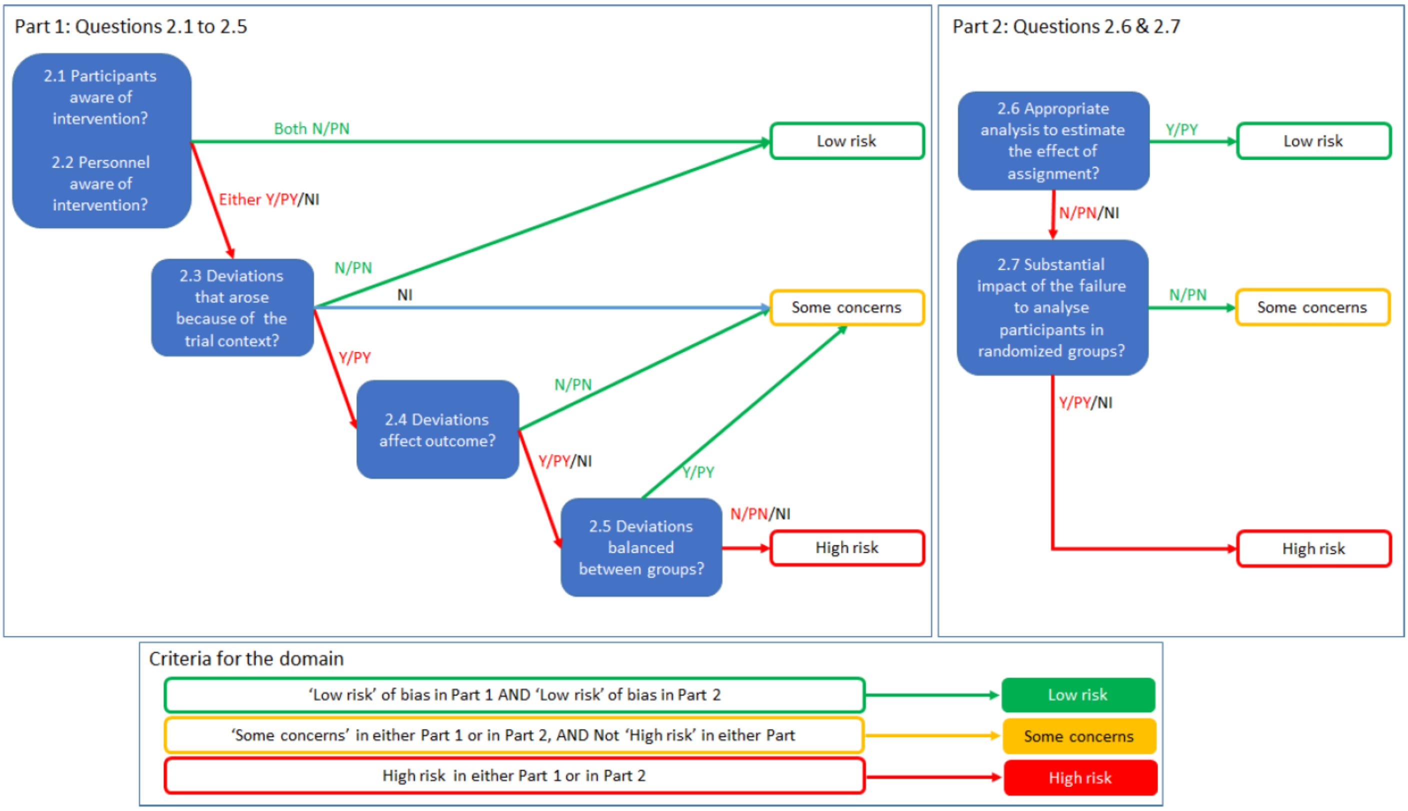


| | |
|---|---|
| 2. Risk of bias judgment: Deviations from intended interventions | ◯ Low<br>◯ High<br>◯ Some concerns<br>(Use above image, if needed, to assist with adjudication based on signaling question response and corresponding bias risk) |
| Include justification for judgment related to deviations from intended interventions here | ____________________<br>(Includes direct quotations from study report text, summary of information from a trial report, etc.) |

#### 3. Bias due to missing outcome data

| | Yes (Y) | Probably Yes (PY) | No (N) | Probably No (PN) | No Information (NI) |
|---|---|---|---|---|---|
| 3.1 Were data for this outcome available for all, or nearly all, participants randomised? | ◯ | ◯ | ◯ | ◯ | ◯ |
| 3.2 Is there evidence that the result was not biased by missing outcome data? | ◯ | ◯ | ◯ | ◯ | ◯ |

| | | | | | |
|---|---|---|---|---|---|
| 3.3 Could missingness in the outcome depend on its true value? | ◯ | ◯ | ◯ | ◯ | ◯ |
| 3.4 Is it likely that missingness in the outcome depended on its true value? | ◯ | ◯ | ◯ | ◯ | ◯ |

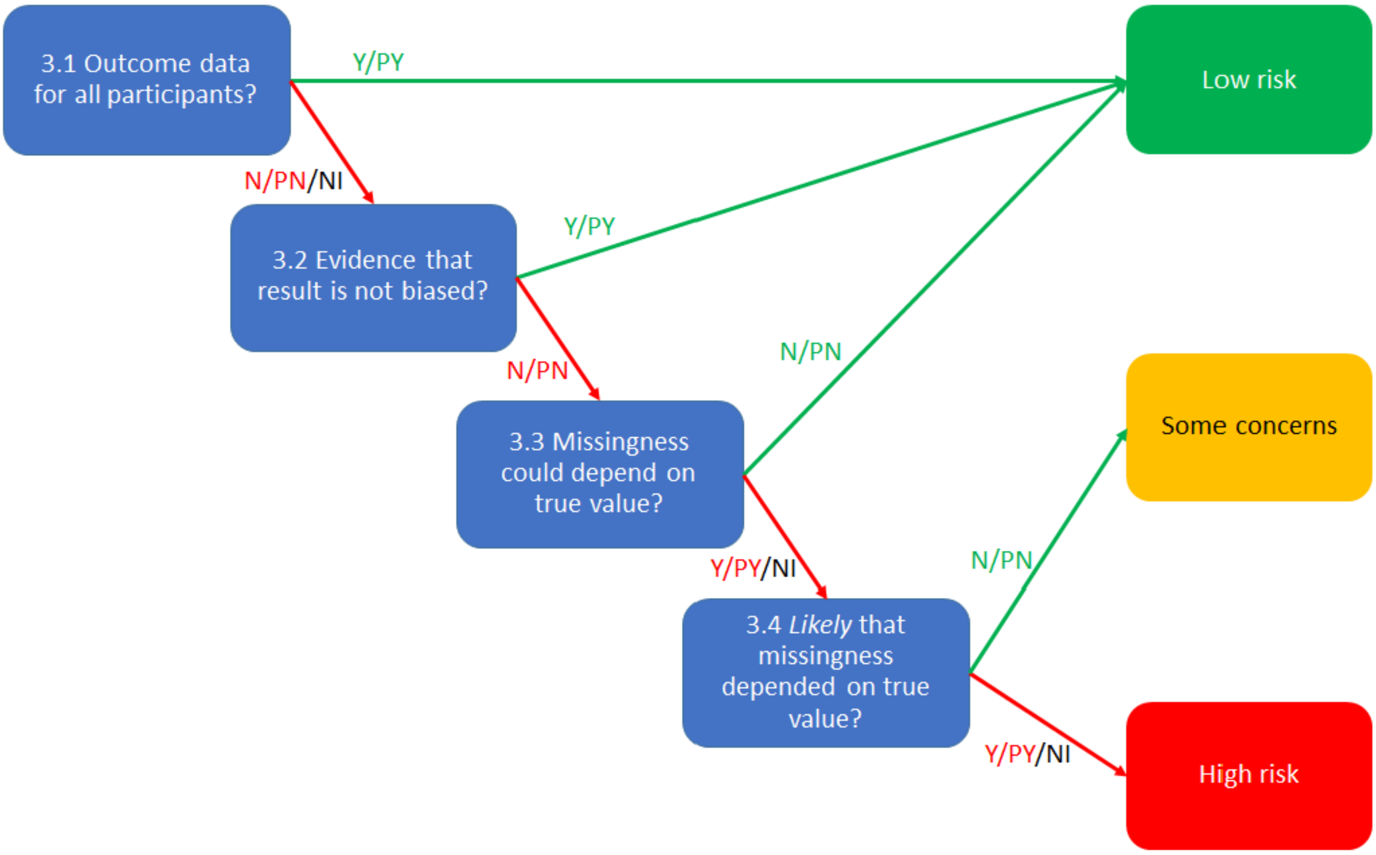


| | |
|---|---|
| 3. Risk of bias judgment: Missing outcome data | ◯ Low<br>◯ High<br>◯ Some concerns<br>(Use above image, if needed, to assist with adjudication based on signaling question response and corresponding bias risk) |
| Include justification for judgment related to missing outcome data here | ______________________________<br>(Includes direct quotations from study report text, summary of information from a trial report, etc.) |

#### 4. Bias in measurement of the outcome

| | Yes (Y) | Probably Yes (PY) | No (N) | Probably No (PN) | No Information (NI) |
|---|---|---|---|---|---|
| 4.1 Was the method of measuring the outcome inappropriate? | ○ | ○ | ○ | ○ | ○ |
| 4.2 Could measurement or ascertainment of the outcome have differed between intervention groups? | ○ | ○ | ○ | ○ | ○ |
| 4.3 Were outcome assessors aware of the intervention received by study participants? | ○ | ○ | ○ | ○ | ○ |
| 4.4 Could assessment of the outcome have been influenced by knowledge of intervention received? | ○ | ○ | ○ | ○ | ○ |
| 4.5 Is it likely that assessment of the outcome was influenced by knowledge of intervention received? | ○ | ○ | ○ | ○ | ○ |

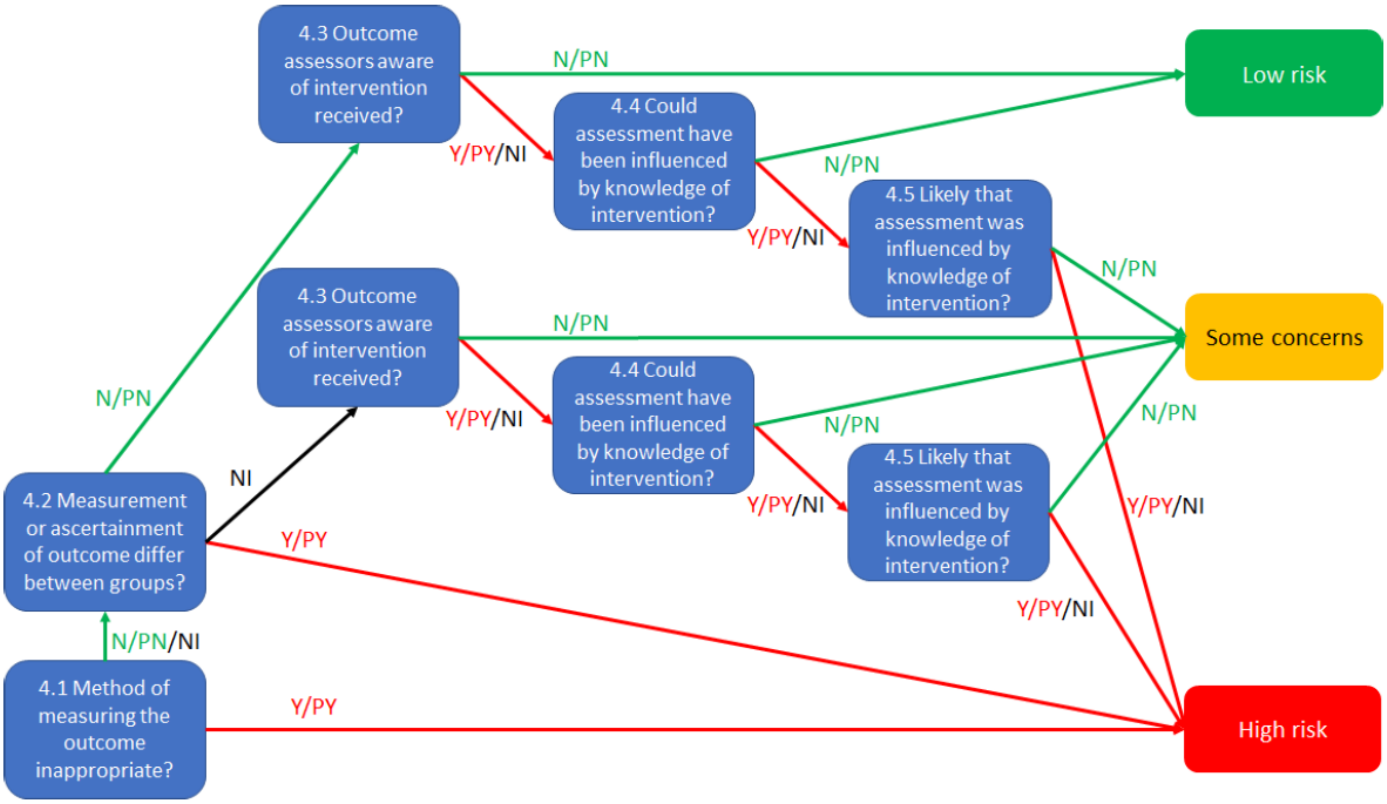

| | |
|---|---|
| 4. Risk of bias judgment: Measurement of the outcome | ◯ Low<br>◯ High<br>◯ Some concerns<br>(Use above image, if needed, to assist with adjudication based on signaling question response and corresponding bias risk) |
| Include justification for judgment related to measurement of the outcome here | ____________________________________<br>(Includes direct quotations from study report text, summary of information from a trial report, etc.) |

#### 5. Bias in selection of the reported result

| | Yes (Y) | Probably Yes (PY) | No (N) | Probably No (PN) | No Information (NI) |
|---|---|---|---|---|---|
| 5.1 Were the data that produced this result analysed in accordance with a prespecified analysis plan that was finalised before unblinded outcome data were available for analysis? | ◯ | ◯ | ◯ | ◯ | ◯ |
| 5.2 Is the numerical result being assessed likely to have been selected, on the basis of the results, from multiple eligible outcome measurements (eg, scales, definitions, time points) within the outcome domain? | ◯ | ◯ | ◯ | ◯ | ◯ |
| 5.3 Is the numerical result being assessed likely to have been selected, on the basis of the results, from multiple eligible analyses of the data? | ◯ | ◯ | ◯ | ◯ | ◯ |

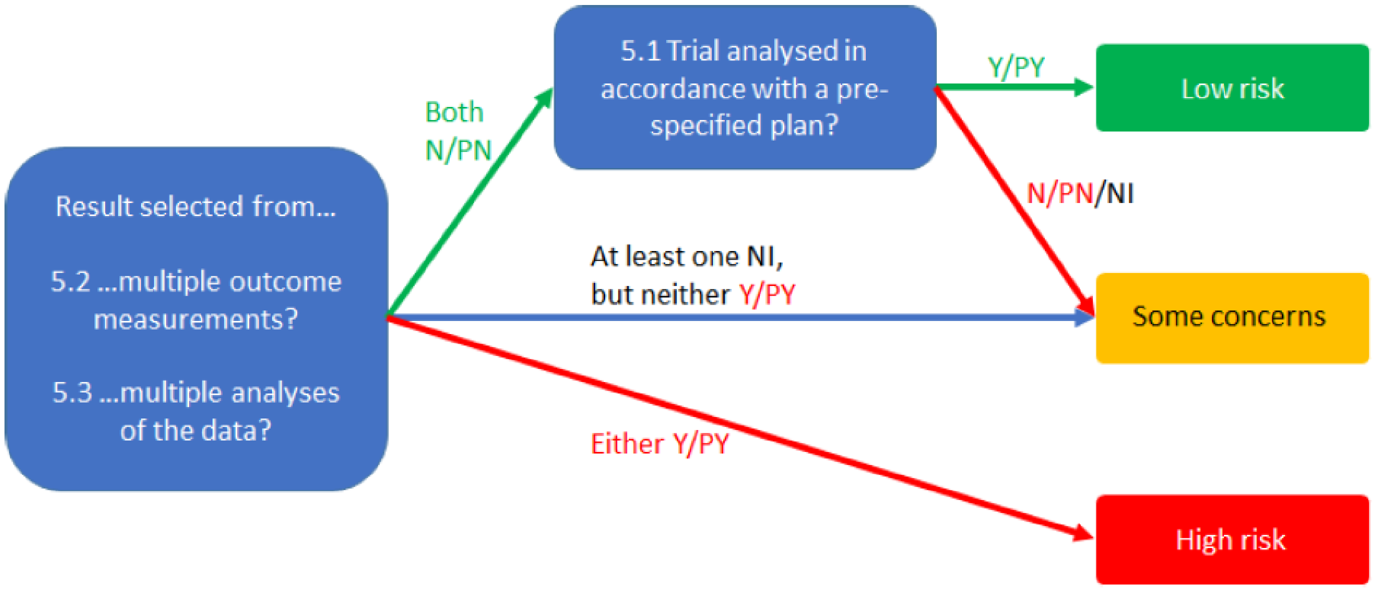


| | |
|---|---|
| 5. Risk of bias judgment: Selection of the reported result | ◯ Low<br>◯ High<br>◯ Some concerns<br>(Use above image, if needed, to assist with adjudication based on signaling question response and corresponding bias risk) |
| Include justification for judgment related to selection of the reported result here | ________________________<br>(Includes direct quotations from study report text, summary of information from a trial report, etc.) |
| Overall risk of bias judgment | ◯ Low<br>◯ High<br>◯ Some concerns<br>(Low = The trial is judged to be at low risk of bias for all domains for this result. Some concerns = The trial is judged to raise some concerns in at least one domain for this result, but not to be at high risk of bias for any domain. High = The trial is judged to be at high risk of bias in at least one domain for this result OR The trial is judged to have some concerns for multiple domains in a way that substantially lowers confidence in the result.) |
| Include justification for judgment related to overall risk of bias here | ________________________ |
